# Bacteriocin production facilitates nosocomial emergence of vancomycin-resistant *Enterococcus faecium*

**DOI:** 10.1101/2024.08.01.24311290

**Authors:** Emma G. Mills, Alexander B. Smith, Marissa P. Griffith, Katharine Hewlett, Lora Pless, Alexander J. Sundermann, Lee H. Harrison, Joseph P. Zackular, Daria Van Tyne

## Abstract

Vancomycin-resistant *Enterococcus faecium* (VREfm) is a prevalent healthcare-acquired pathogen. Gastrointestinal colonization can lead to difficult-to-treat bloodstream infections with high mortality rates. Prior studies have investigated VREfm population structure within healthcare centers. However, little is known about how and why hospital-adapted VREfm populations change over time. We sequenced 710 healthcare-associated VREfm clinical isolates from 2017-2022 from a large tertiary care center as part of the Enhanced Detection System for Healthcare-Associated Transmission (EDS-HAT) program. Although the VREfm population in our center was polyclonal, 46% of isolates formed genetically related clusters, suggesting a high transmission rate. We compared our collection to 15,631 publicly available VREfm genomes spanning 20 years. Our findings describe a drastic shift in lineage replacement within nosocomial VREfm populations at both the local and global level. Functional and genomic analysis revealed, antimicrobial peptide, bacteriocin T8 may be a driving feature of strain emergence and persistence in the hospital setting.

**Summary:** This study shows local and global lineage replacement of vancomycin-resistant *Enterococcus faecium*. Bacteriocin T8 is enriched in emergent lineages and provides a strong competitive advantage *in vitro* and *in vivo*.

## INTRODUCTION

*Enterococcus faecium* is a gastrointestinal tract commensal that can also cause serious infections, most commonly bloodstream and urinary tract infections, especially in immunocompromised and hospitalized patients^1^. Hospitalized patients are often exposed to high levels of antibiotics, which decrease the diversity of commensals in the GI tract and facilitate the overgrowth of multidrug-resistant organisms like vancomycin-resistant *E. faecium* (VREfm)^2–4^. VREfm overgrowth within the intestinal tract predisposes patients to invasive bloodstream infections^2,4–7^. Further, increased VREfm GI tract burdens cause patients to shed VREfm into the environment, facilitating transmission to other patients mainly through the fecal-oral route^8–10^.

Whole genome sequencing facilitates the surveillance and characterization of VREfm population structure and transmission dynamics within healthcare settings. Multi-locus sequence typing (ST) allows tracking of VREfm lineages both within and between healthcare facilities and on both local and global scales^11–13^. STs with similar genotypes, defined as having 4 or more identical loci, can be grouped into clonal complexes. VREfm lineages most often belong to clonal complex 17 (CC17), which phylogenetically resides within hospital-adapted *E. faecium* Clade A1. CC17 strains frequently encode antimicrobial resistance genes, mobile genetic elements, and genes that enable the metabolism of amino sugars found on GI epithelia and mucin, likely contributing to the success of CC17 strains in healthcare settings^12–18^. This success is exemplified by CC17 lineages being identified as responsible for widespread outbreaks and increased rates of invasive infection^14,15,17,19^. Although several prior studies have investigated VREfm population structure and dynamics within healthcare settings, we know little about the factors that drive the emergence and persistence of particular VREfm lineages in the hospital.

In this study, we characterized the population structure and dynamics of VREfm within a single hospital using whole genome sequencing-based surveillance and functional characterization of genes associated with nosocomial emergence. We systematically collected 710 VREfm clinical isolates over a 6-year period and used both genomic analysis and phenotypic testing to investigate factors contributing to population shifts observed within the facility. Additionally, we compared local findings with a global collection of 15,631 publicly available VREfm genomes isolated from human sources from 2002-2022. We found that a bacteriocin produced by some VREfm lineages provided a strong competitive advantage, highlighting an adaptive mechanism that likely contributes to lineage replacement of VREfm on both local and global scales.

## RESULTS

### Population structure and genomic epidemiology of VREfm at a single hospital

Between 2017 and 2022, the Enhanced Detection System for Hospital-Associated Transmission (EDS-HAT) whole genome sequencing surveillance program collected 710 healthcare-associated VREfm isolates, *i.e.* isolates collected from patients with hospital stays >2 days or prior 30-day healthcare exposures at UPMC. The most common isolate sources were urine (42%), blood (24%), and wound sites (19%) (**Supplementary Table 1**). We first investigated the genomic diversity of this collection through multi-locus sequence typing (ST), which identified 42 different STs. All isolates belonged to hospital-adapted lineages within CC17, including ST17 (23%), ST117 (13%), ST1471 (11%), and ST80 (10%) (**Fig. 1A, Supplementary Table 1,**). To characterize the population structure of our collection, a core-genome phylogenetic tree was constructed based on 1604 core genes (**Fig. S1A**). Despite being entirely comprised of CC17 strains, the VREfm population displayed variable genetic diversity within and between STs and showed evidence that some isolates were closely related to one another. To assess genetic relatedness among the collected isolates, we performed split kmer analysis (SKA) to cluster isolates that had fewer than 10 single nucleotide polymorphisms (SNPs) in pairwise comparisons. This analysis revealed 112 putative transmission clusters that contained 2-9 isolates and encompassed 46% of the collection (**Fig. 1A**). Despite a high degree of clustering among all isolates, the proportion of isolates residing in putative transmission clusters was variable between STs. Although ST17 was the most prevalent lineage, it had the lowest percentage of clustered isolates (33%, 54/165 isolates, *p* < 0.001). On the other hand, ST1478 showed a significantly higher percentage of clustered isolates (69%, 38/55 isolates, *p* <0.004) (**Fig. 1B**).

**Figure 1:**
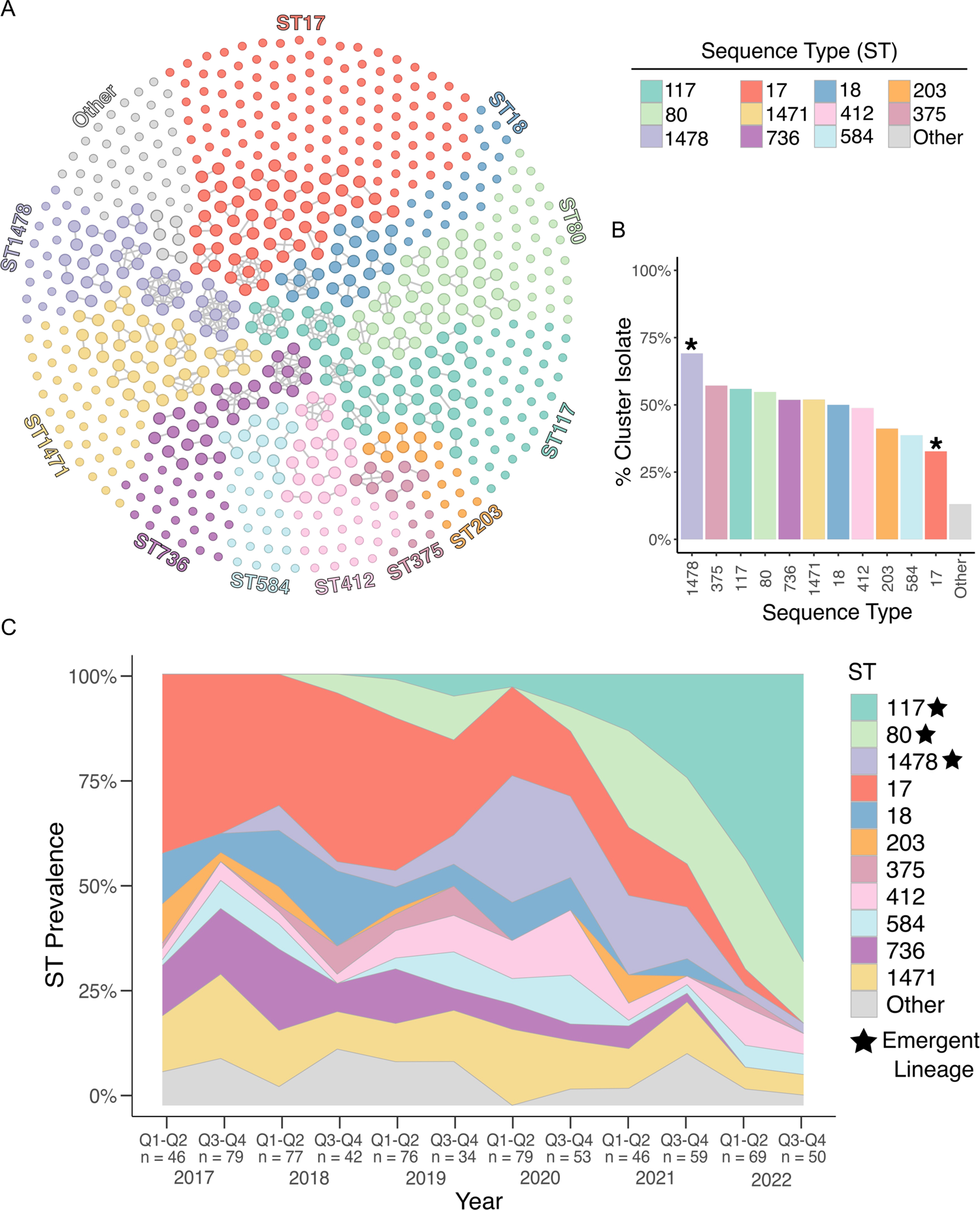
Population structure and temporal dynamics of VREfm at UPMC over 6 years. (A) Cluster network diagram of 710 sequenced VREfm genomes constructed using Gephi v0.10. Isolates are grouped and colored by multilocus sequence type (ST). Isolates that fall within putative transmission clusters (≤ 10 SNPs) are connected with grey lines. (B) Prevalence of cluster isolates within different STs. Asterisks mark STs that show a higher or lower percentage of clustered isolates compared to the total collection (p < 0.004). (C) Biannual distribution of STs. Q1-Q2 = January-June; Q3-Q4 = July-December. The number of samples within each sample period is noted below.

### VREfm lineage replacement

We next investigated how the VREfm population changed over time within UPMC by characterizing the ST distribution over the collection period in 6-month intervals (**Fig. 1C**). Prior to 2020, ST17 was the most frequently sampled ST, making up 34% of the collection between 2017 and 2019. However, during 2020, the emergence of ST1478 (23%) coincided with the decline of ST17 (17%). For the remainder of the collection period, the presence of ST17 continued to decline, and this lineage was not detected during the second half of 2022 (**Fig. 1C**). In contrast, lineages ST80 and ST117 were not detected in 2017, but together rose to 81% by the end of 2022, effectively replacing ST17 and other lineages that were previously detected. We therefore designated ST80, ST117, and ST1478 as emergent lineages at UPMC.

To identify factors contributing to lineage replacement, we first investigated the frequency of non-susceptibility to the clinically relevant antibiotics linezolid and daptomycin (**Fig. S1, Supplementary Table 1**). The emergent lineages (ST80, ST117, ST1478) did not show a higher frequency of non-susceptible isolates, with non-susceptibility rates of 0-5% (linezolid) and 6-20% (daptomycin). We then investigated the distribution of genomic features such as antimicrobial resistance genes (ARGs), virulence factors, and plasmid replicons among the different lineages (**Fig. S1C-E**). We observed variation in the number of ARGs and virulence genes within the emergent lineages, with ST117 (mean = 15.6 ARGs) and ST1478 (mean = 16.0 ARGs) having more ARGs compared with ST17 (mean = 14.1, *p* <0.0001) (**Fig. S2C**). The macrolide efflux transporter *mefH* was found in nearly all ST117 and ST1478 isolates and was only identified in 4 other isolates (**Supplementary Table 2**). Similarly, the aminoglycoside nucleotidyltransferase *ant(6)-Ia* was highly enriched in these two lineages, being present in 99% and 93% of ST117 and ST1478 isolates, compared with 52% of other isolates. ST117 also had more virulence genes (mean = 4.0) compared to ST17 (mean = 3.7, *p* < 0.0001) (**Fig. S2D**). Virulence genes enriched (>98%) in ST117 genomes included the colonization factors *acm, fss3, ecbA,* and *sgrA* (**Supplementary Table 2**). ST1478 and ST117 genomes also encoded more plasmid replicons compared with the historical lineage ST17 (*p* < 0.0001) (**Fig. S2E**). We further investigated the distribution of replicons among lineages and found the rep11a replicon was present at higher frequency in emergent lineages ST80 (64%), ST117 (75%), and ST1478 (95%), versus only 15% of other isolates (**Supplementary Table 2**). Similarly, the repUS15_2 family replicon was enriched in ST117 (95%) and ST1478 (98%) but was seen at a lower prevalence the remaining isolates (28%), including ST80 (10%) (**Supplementary Table 2**). Together these data suggest that emergent lineages possess genomic features that might facilitate their emergence within the hospital.

### Growth inhibition caused by emergent isolates is associated with bacteriocin T8

To determine other factors contributing to lineage replacement, we investigated whether emergent lineage VREfm isolates inhibited the *in vitro* growth of historical lineage isolates. We first performed a pairwise spot killing assay using the earliest available isolates from the historical lineage ST17 and the emergent lineage ST117. We found that the ST117 isolate was able to inhibit growth of the historical ST17 isolate, causing a large zone of inhibition in the ST17 isolate lawn surrounding the ST117 isolate spot (**Fig. 2A**). We then conducted pairwise spot killing assays using isolates from each of the 11 lineages having ≥ 10 isolates in the dataset (**Fig. 2B, Supplementary Table 3**). We found that isolates from all three emergent lineages caused growth inhibition of isolates from other lineages. Bacteriocins are antimicrobial peptides which have been widely studied in *Enterococcus* due to their ability to inhibit growth of other bacteria, and their potential role as probiotics^20–23^. Therefore, we screened the genomes in the dataset for predicted bacteriocins and found one bacteriocin, called T8, that was differentially present and found in 36% of isolates (**Fig. S2A, Supplementary Table 2**).

**Figure 2:**
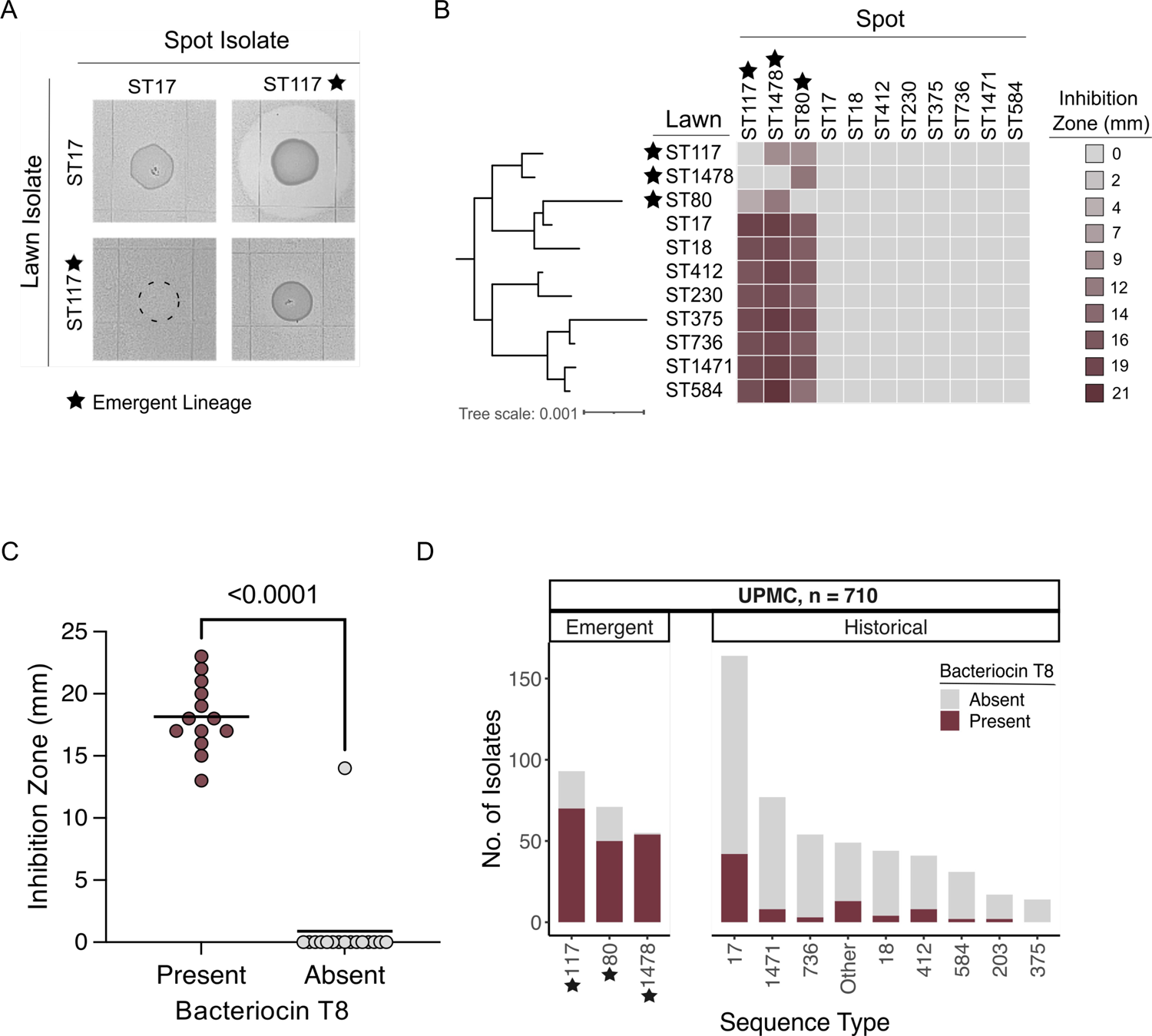
Bacteriocin T8 is associated with growth inhibition in emergent lineage isolates. (A) Pairwise spot killing assay with reference ST17 (historical) and ST117 (emergent) isolates. Dashed circle shows where the ST17 isolate was spotted onto the ST117 lawn but did not grow. (B) Pairwise spot killing assay with reference isolates from each of the 11 main lineages within the UPMC collection. Inhibition zone quantification (mm) is shaded from most inhibition (burgundy) to no inhibition (grey). Inhibition zone values were averaged from three biological replicates. The midpoint-rooted phylogenetic tree was constructed using RAxML HPC with 100 bootstraps based on a core genome alignment produced by Roary. (C) Spot killing assay results of 28 VREfm isolates spotted onto a lawn of the bacteriocin T8-negative, ST17 reference isolate. Isolates are grouped by presence (burgundy) or absence (grey) of bacteriocin T8 in their genome. P-value indicates significance from a two-tailed Mann-Whitney test with α = 0.05. (D) Abundance of bacteriocin T8 within main STs at UPMC. Emergent lineages are noted with a black star.

Bacteriocin T8 is identical to two other enterococcal bacteriocins, named hiracin JM79^21^ and bacteriocin 43^22^, and all three names have been used in various prior studies to describe what is now known to be the same bacteriocin. We chose to refer to this bacteriocin as T8 because this is how it was first and most frequently described in the literature. To quantify the association of bacteriocin T8 with growth inhibition, we screened 28 VREfm isolates representing 11 STs against the same ST17 reference isolate to assess growth inhibition, and found that bacteriocin T8 presence was strongly associated with growth inhibition (*p* < 0.0001) (**Fig. 2C** and **Supplementary Table 4)**. A single isolate, called VRE36503, lacked bacteriocin T8 but still showed growth inhibition of the ST17 lawn. While no predicted bacteriocins were identified in the VRE36503 genome using the BAGEL4 prediction tool, an additional search for secondary metabolites identified a gene cluster with homology to the carnobacteriocin XY biosynthetic gene cluster^24^, suggesting that growth inhibition by VRE36503 was independent of bacteriocin T8.

To investigate whether bacteriocin T8 was encoded by a plasmid, we performed long-read sequencing and hybrid genome assembly on a bacteriocin T8-positive ST117 isolate. We found that bacteriocin T8 and the corresponding immunity factor were carried on a 6,173 bp rep11a-family plasmid that also encoded mobilization genes *mobABC,* allowing for plasmid transfer (**Fig. S2B**). Of all bacteriocin T8-positive isolates (n = 253), rep11a was found in 93% based on short-read assembly data. We also characterized the distribution of bacteriocin T8 among the main lineages in the dataset, and identified a high prevalence of bacteriocin T8 isolates among the emergent lineages ST80 (70%), ST117 (74%), and ST1478 (96%) (**Fig. 2D**). Bacteriocin T8 was only found in 16% of the remaining isolates in the collection, most of which belonged to ST17 (25%). Due to the enrichment of bacteriocin T8 in emergent lineage genomes, we hypothesized that it might provide a competitive advantage to VREfm during colonization and infection of hospitalized patients.

### Bacteriocin T8 expression provides a competitive advantage *in vitro* and *in vivo*

We confirmed that bacteriocin T8 caused growth inhibition by transforming a bacteriocin T8-negative, clinical *E. faecium* isolate (referred to as Parent) with pBAC (plasmid containing bacteriocin T8 and immunity factor) or pEV (empty vector). To test whether pBAC conferred growth inhibition, we performed a pairwise spot killing assay and found that the pBAC strain caused a large zone of inhibition on a lawn of the pEV strain (**Fig. 3A**). Next, we quantified the competitive advantage of the pBAC strain by performing a liquid competition assay. We independently competed the pBAC and pEV strains against the Parent strain at 50:50 and 10:90 starting ratios and quantified the abundance of each strain in the mixture after 24 and 48 hours. At both ratios and timepoints, the pBAC strain was able to outcompete the Parent strain to a much greater extent compared with the pEV strain (p < 0.01) (**Fig 3B, Supplementary Table 5**). We then evaluated if the competitive advantage conferred by bacteriocin T8 *in vitro* translated to the mammalian gut. To assess this, we pretreated C57BL/6 mice with vancomycin to deplete their endogenous *Enterococcus* population before orally gavaging mice with the pBAC or pEV strains for two days. We monitored the abundance of each strain in stool for eight days following the initiation of infection (**Fig 3C, Supplementary Table 6**). On Day 1 there was no difference in GI burden between the two groups, indicating that mice received similar inocula of each strain. However, at all subsequent time points the pBAC strain was detected at a significantly higher abundance compared to the pEV strain (*p* < 0.05) (**Fig. 3C**). These findings suggest that bacteriocin T8 provides a competitive advantage to VREfm in the mammalian GI tract.

**Figure 3:**
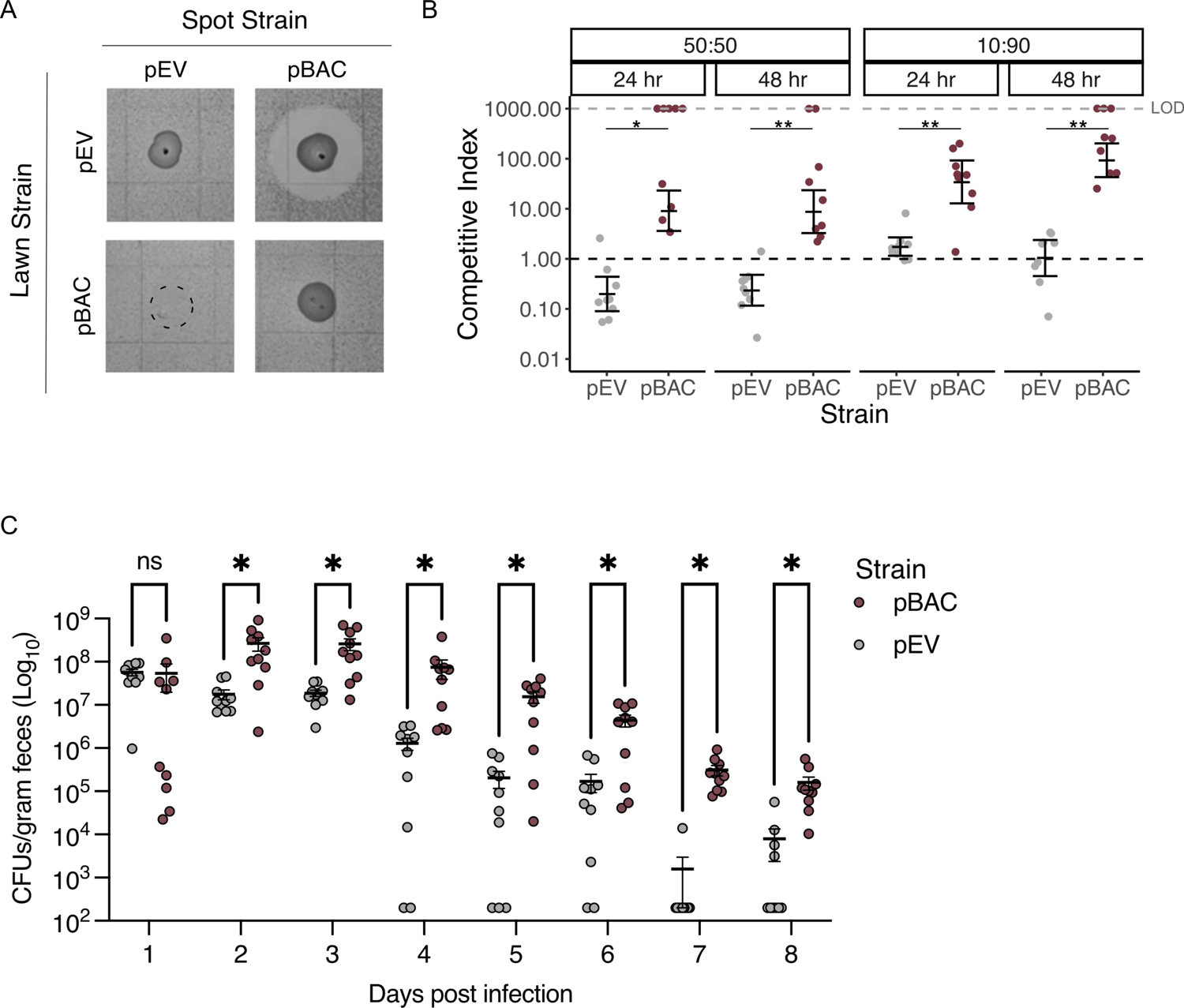
Bacteriocin T8 production provides a competitive advantage *in vitro* and increases *E. faecium* colonization *in vivo.* (A) Pairwise spot-killing assay of pBAC and pEV strains. Dashed circle shows where the pEV strain was spotted onto the pBAC lawn but did not grow. (B) Liquid competition assay. pBAC and pEV were independently competed against the parent strain at 50:50 and 10:90 ratios. Samples were taken after 24 and 48hrs to calculate CFU/mL. Assays were performed in 3 technical replicates each consisting of 3 biological replicates. Instances where the parent strain fell below the limit of detection (LOD, shown with grey dashed line) were not included in statical analyses. The distribution of competitive index at each time point and ratio were compared between pBAC and pEV using a two-tailed Mann Whitey test, (\**p <* 0.01, \*\**p* < 0.001). (C) Colonization of pBAC and pEV strains in the murine gut. Mice were orally gavaged with either pBAC or pEV for 2 days. Stool samples were collected starting on Day 1 after initiation of infection to quantify CFU/g of each strain over time. A two-tailed Mann Whitney test was used assess CFU/g distribution between pBAC and pEV strains (\**p <* 0.04).

### Bacteriocin presence is associated with global VREfm lineage replacement

We next sought to determine if the lineage replacement we observed at UPMC was reflective of global VREfm population dynamics. To investigate this question, we gathered 15,631 publicly available VREfm genomes collected from human sources between 2002-2022 (**Supplementary Table 7**). This collection consisted of genomes from 53 countries; however, the majority of isolates were from the United States (23%), Denmark (23%), and Australia (20%) (**Fig. S3, Supplementary Table 7**). To investigate VREfm global genomic diversity, we performed sequence typing on this collection and examined the distributions of STs by continent (**Fig. 4A**). Europe and Asia had relatively clonal populations, while the populations in North America and Australia were more diverse. ST80 was the single most prevalent ST (20%) and was mainly found in Europe (30%) and Asia (36%). ST117 was the second most prevalent ST (12%), and had the highest prevalence in Europe (18%) and North America (12%). ST17 accounted for 7% of the global population and was sampled predominantly in North America (18%). To characterize global population dynamics of VREfm, we investigated the prevalence of STs over the 20-year global collection period (**Fig. 4B, Supplementary Table 7**). Prior to 2010, ST17 and ST18 were the most prevalent lineages, while ST80 and ST117 were detected very infrequently. After 2010, however, ST117 and ST80 rose to 60% by the end of 2022, effectively replacing ST17 (3%) and ST18 (0.2%). These data suggest that the emergence of ST80 and ST117 that we observed locally was reflective of global trends.

**Figure 4:**
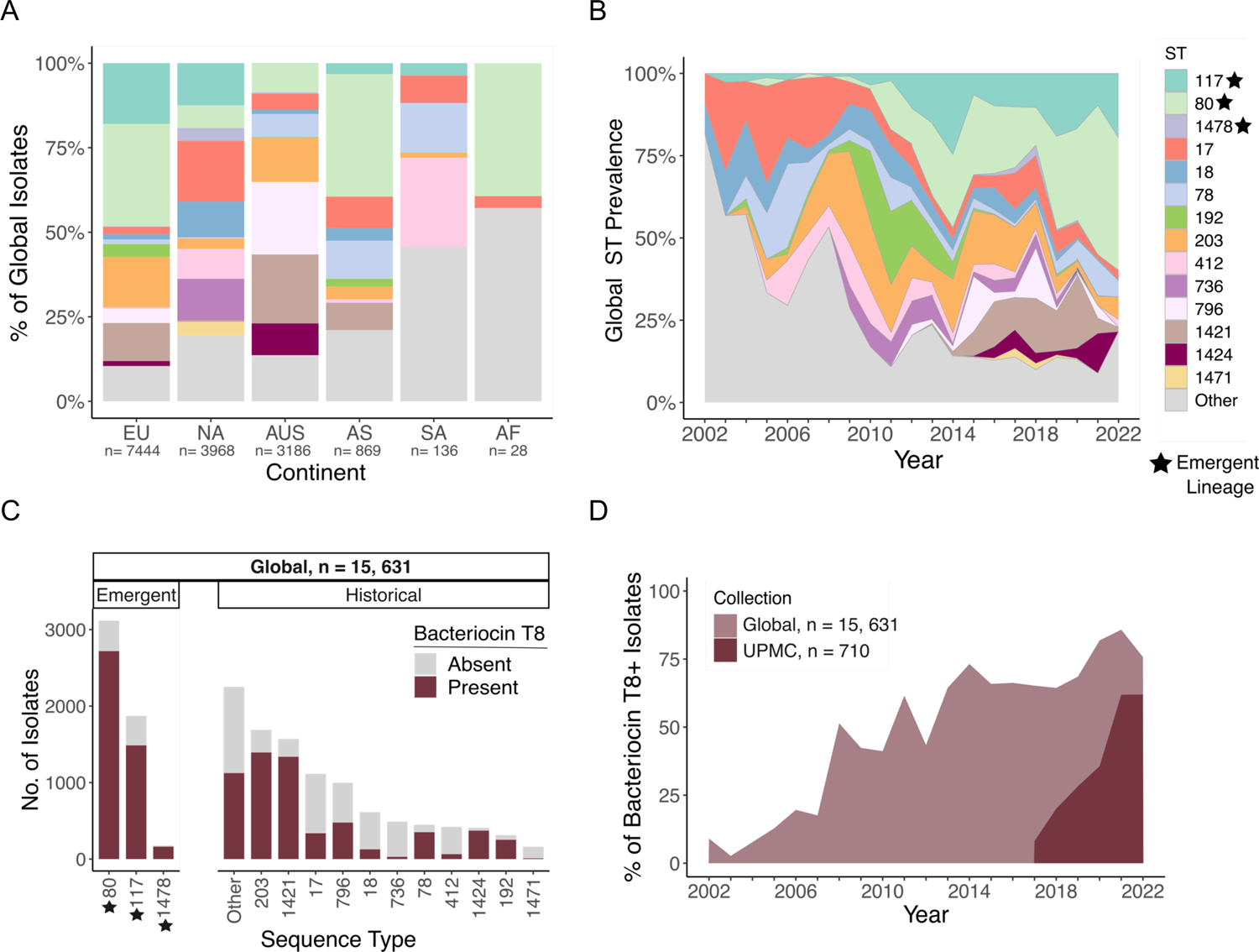
Global bacteriocin T8 prevalence increases over time and is associated with emergent lineages. (A) Geographic distribution of 15,631 global VREfm isolates across continents; EU: Europe, NA: North America, AUS: Australia, AS: Asia, and AF: Africa. Isolates are colored by sequence type (ST). (B) Global ST distribution between 2002-2022. Isolates are colored by ST. (C) Abundance of bacteriocin T8 within main STs. Bars are separated into emergent and historical lineages based on trends seen within the UPMC collection (Fig. S3). Bacteriocin T8 presence is shaded in burgundy and absence in grey. (D) Prevalence of bacteriocin T8 over time at UPMC (burgundy) and in the global collection (light burgundy).

To investigate if bacteriocin T8 was similarly enriched in emergent lineages, we examined the distribution of bacteriocin T8 among the STs sampled in the global collection of VREfm isolated from human sources (**Fig. 4C**). Bacteriocin T8 was enriched in emergent lineages ST80, ST117, and ST1478 globally, with more than 79% of isolates in each ST predicted to encode the bacteriocin. Similar to our local prevalence (25%), bacteriocin T8 was found in only 30% of isolates in the previously dominant lineage ST17. We also investigated if bacteriocin T8 was increasing over time within both collections (**Fig. 4D**). Within the local UPMC collection, bacteriocin T8 presence rose from 8% in 2017 to 62% in 2022. Similarly, we observed a 67% increase in the prevalence of bacteriocin T8 between 2002 and 2022 in the global collection. Within both collections, the increase in bacteriocin T8 prevalence was associated with the replacement of the historical ST17 lineage with emergent lineages ST80 and ST117. Taken together, these findings suggest that bacteriocin T8 may be a driving feature of global VREfm strain emergence and persistence in healthcare settings.

## DISCUSSION

In this study, we examined the population structure and dynamics of 710 VREfm clinical isolates collected over 6 years from a single hospital. A significant strength of our study lies in the use of a systematic collection of hospital-acquired VREfm isolates collected over a multi-year period. Additionally, through comparing our findings to a large global collection of over 15,000 VREfm isolates from human sources, we confirmed that many of our findings were generalizable to other settings worldwide. Our data show the emergence of ST80 and ST117 both locally and globally, highlighting the strong competitive advantage of these lineages and identifying bacteriocin T8 as a likely contributor to VREfm lineage replacement.

Similar to other studies, we found that the VREfm population at our hospital was polyclonal, with the majority (57%) of isolates belonging to 4 prevalent lineages: ST17, ST117, ST80, and ST1471^12,13,25,26^. These lineages belong to the hospital-adapted Clade A1 of *E. faecium* and also belong to CC17, which is known to be highly epidemic within healthcare systems and the cause of widespread outbreaks^13–15,27,28^. While previous studies have reported nosocomial VREfm transmission rates ranging from 60-80%^28–31^, we found that only 46% of isolates in our dataset belonged to putative transmission clusters. This difference is likely due to our use of a 10 SNP cut-off for clustering and not including sampling of VREfm from GI tract colonization, which might limit our ability to detect transmission^32^. We did however note differences in the percentage of isolates residing within putative transmission clusters among different lineages, with lineage ST1478 showing a significantly higher proportion of clustered isolates. Prior literature has found that the ST117 lineage was responsible for numerous VREfm hospital outbreaks^12,14,33^; however, the ST1478 lineage has only been detected in the US and Canada^19^. Taken together, these findings suggest that some VREfm lineages might be more efficient than others at transmitting between patients in the hospital.

Through phenotypic screening and comparative genomic analysis, we found that the antimicrobial peptide bacteriocin T8 was enriched in emergent lineages both locally and globally, and that it conferred a growth advantage to *E. faecium* both *in vitro* and *in vivo*. The enrichment of bacteriocin T8 in emergent lineages and increasing prevalence over time suggests that acquisition of this bacteriocin is highly advantageous. Bacteriocins have been shown to facilitate expansion of bacterial populations by killing susceptible bacteria, thereby carving out a stable environment for the expansion of bacteriocin-expressing bacteria^34–39^. A prior study by Kommineni et al. investigated the competitive advantage conferred by bactericion-21 to *E. faecalis* in the mammalian GI tract^34^. Similar to our findings, bacteriocin production in that study was associated with increased GI tract colonization in the murine gut. Further, the study found that the production of bacteriocin-21 was able to clear a vancomycin-resistant *E. faecalis* strain from the GI tract^34^. Due to the strong inhibitory activity of bacteriocins, they are an attractive avenue for development as new antimicrobial interventions, such as inclusion in probiotics and food preservation^36,40^. A recent study showed that a genetically engineered probiotic *E. coli* strain containing 3 bacteriocins, including bacteriocin T8 (referred to as hirJM79), was able to clear vancomycin-resistant *E. faecium* and *E. faecalis* in a murine model of enterococcal colonization^36^. Although this result is exciting, it is somewhat troubling that based on our global findings the vast majority of VREfm isolates sequenced in 2022 already encoded bacteriocin T8 and the associated immunity gene, suggesting they would be resistant to bacteriocin T8 activity.

Our study had several limitations. First, within our UPMC collection we only investigated VREfm isolates collected from clinical infections that were suspected to be hospital acquired. Isolates not meeting inclusion criteria, including potentially community-acquired VREfm, were not included. We also very likely under-sampled the full VREfm population diversity within our center because many hospitalized patients have asymptomatic GI tract colonization^32^. The global collection we analyzed was biased towards countries with high rates of VREfm infection and with the infrastructure and capacity to perform high-throughput sequencing, which resulted in some continents, like Asia and Africa, to be greatly under-sampled. Secondly, we focused on bacteriocin T8 as a contributor to lineage success; however, this may not be the only factor driving the lineage replacement that we observed. We did not investigate other potential adaptations among emergent lineages, such as virulence factors that were enriched in the ST117 lineage. Further, it is important to note that *E. faecium* virulence factors in general are undercharacterized, limiting their identification across our collection. Moreover, additional uncharacterized mutations within the emergent lineages could contribute to antimicrobial resistance or tolerance, potentially aiding in lineage replacement. Additionally, in the murine model we focused on the impact of bacteriocin production on enterococci, potentially overlooking other microbiome disruptions that could be explored through additional metagenomic sequencing.

In summary, we characterized the local and global population structure and temporal dynamics of VREfm using comparative genomics and functional analyses. Through investigating VREfm populations sampled over 6 years at our healthcare center and over 20 years globally, we identified lineage replacement associated with the spread of strains encoding bacteriocin T8. Phenotypic characterization showed that bacteriocin T8 likely contributes to VREfm lineage replacement by conferring a strong competitive advantage that is observed both *in vitro* and *in vivo.* Although we identified bacteriocin T8 production as a potential adaptive mechanism directing VREfm lineage replacement, this study prompts further investigation into other features driving the evolutionary dynamics in this important and difficult to treat pathogen.

## METHODS

### Study Setting

This was a retrospective observational study of VREfm collected from patients at the University of Pittsburgh Medical Center (UPMC) by the Enhanced Detection System for Healthcare-Associated Transmission (EDS-HAT)^41^. UPMC is an adult tertiary care hospital with 699 beds (including 134 critical care beds) and performs >400 solid organ transplants each year. A total of 710 VREfm clinical isolates were collected from patients with a hospital admission date ≥ 2 days prior or with a recent healthcare exposure within 30-days before the culture date, from January 2017 to December 2022. Available daptomycin and linezolid minimal inhibitory concentration (MIC) data was collected from patient records and interpreted using Clinical & Laboratory Standards Institute (CLSI) M100 guidelines. The study was approved in its entirety by the institutional review board at the University of Pittsburgh (STUDY21040126).

### Whole genome sequencing (WGS) and bioinformatic analyses

Genomic DNA was extracted using a DNeasy Blood and Tissue Kit (Qiagen, Hilden, Germany) from VREfm isolates that were grown overnight at 37°C on blood agar plates. Following DNA extraction, next-generation sequencing libraries were generated using the Illumina DNA Prep protocol and then sequenced (2×150 bp, paired-end) on a NextSeq500, NextSeq2000, or MiSeq. The resulting reads were assembled using SPAdes v3.15.5^42^. Assembly quality was determined using QUAST v5.2.0^43^. Assemblies passed quality control if the coverage was > 35X and the assembly had < 350 contigs. Species were identified and possible contamination was detected using Kraken2 (v2.0.8-β)^44^. Multilocus sequence types (STs) were identified using the PubMLST database with mlst v2.11^45,46^. Isolates with undefined STs were uploaded to the PubMLST server and if their ST was a single locus variant (SLV) of a known ST, they were grouped with the latter. Clusters of genetically related isolates were identified using split kmer analysis v1.0 (SKA)^47^ with average linkage clustering and a 10 SNP cut-off. Genomes were annotated using PROKKA v1.14.5^48^. A cluster network diagram was visualized using Gephi v0.10 with the Fruchterman-Reingold layout^49^. Phylogenetic trees were built using core genome alignments produced by Roary v3.13.0^50^. Gaps in the core genome alignment were masked using Geneious (Geneious Biologics 2024, https://www.geneious.com/biopharma).

Trees were constructed using RAxML HPC v8.2.12 with 100 bootstraps^51^. In the UPMC collection, bacteriocins were identified using BAGEL4 with ≥ 95% coverage and identity^52^. antiSMASH v7.1.0 was used to further identify secondary metabolites in the VRE36503 genome^53^. To identify bacteriocin T8 in the global genome collection, a custom database consisting of the nucleotide sequence for bacteriocin T8 and the corresponding immunity factor was built using ABRicate, and gene presence was defined as hits with ≥ 95% coverage and identity^54^. Antimicrobial resistance genes were identified with AMRFinderPlus v3.12.8^55^.

Presence of plasmid replicons and virulence factors were determined using ABRicate v1.0.01^54^ with the PlasmidFinder^56^ and VFDB^57^ databases, respectively. Gene presence was defined as hits with ≥ 90% coverage and identity. The bacteriocin T8-encoding plasmid was resolved using Unicycler v0.5.0^58^ to hybrid assemble Illumina and MinION (MinION device with R9.4.1 flow cells (Oxford Nanopore Technologies, Oxford, UK) sequencing data collected from isolate VRE38098.

### Spot Killing Assay

Test strains were cultivated for 16-18 hours at 37°C in Brain Heart Infusion (BHI) broth. Subsequently, 5 mL of a BHI top agar lawn (containing 0.35% agar) was prepared by mixing the molten agar with 100 μl of a 1:100 dilution of the overnight culture. This mixture was poured on top of 25 mL of solid BHI agar. Competing strains were spotted (5 μl undiluted overnight culture) onto solidified top agar lawns. Inhibition zones were measured (in mm) after 16-18 hours incubation at 37°C.

### Cloning and expression of Bacteriocin T8

Bacteriocin T8 and the corresponding immunity factor were cloned into the nisin-inducible expression vector pMSP3535, which was modified to encode the chloramphenicol resistance gene *cat* as a positive selectable maker^59^. The insert sequence was amplified from VRE38098 genomic DNA by PCR using primers 5’-AGA CCG GCC TCG AGT CTA GAA TGG GAC TGA TGA ATC AGA ATTG-3’ and 5’-GCG AGC TCG TCGACA GCG CTC AGG CGT TAC TTG GTA GTA TAC-3’. The vector was amplified by PCR using primers 5’-AGC GCT GTC GAC GAG CTC GCAT-3’ and 5’ TCT AGA CTC GAG GCC GGTCTCC-3’. Amplified insert and vector were purified using a PCR Purification Kit (Qiagen), and Gibson assembly was conducted using a HiFi DNA Assembly Cloning Kit (New England Biolabs)^60^. The Gibson product was then transformed into NEB 5-alpha competent *E. coli*, and transformants were selected on BHI agar containing 10 µg/mL chloramphenicol. Plasmids were amplified in 200 ml cultures, harvested by Maxi prep, and sequenced to confirm their identity. The bacteriocin T8-encoding vector (pBAC) or pMSP3535 empty vector (pEV) were then transformed into the bacteriocin T8-negative *E. faecium* ST412 strain DVT705, which is a vancomycin-susceptible derivative of the 14-10-S strain^7^. Successful transformation was confirmed with PCR using pMSP3535 backbone-specific primers 5’-CAA TAC GCA AAC CGC CTCTC-3’ and 5’-TGG CAC TCG GCA CTT AATGG-3’. Inhibitory activity of DVT705 transformed with pBAC was confirmed by a pairwise spot killing assay against DVT705 transformed with pEV.

### Liquid Competition Assay

pBAC and pEV strains were competed individually against the parent DVT705 strain at two ratios, 50:50 and 10:90. For each ratio, three technical replicates each consisting of three biological replicates were performed. Prior to competition, each strain was grown separately overnight and cultures were normalized to OD_600_ = 0.5. Strains were mixed together at the above starting ratios and the mixture was then diluted 1:100 into 5 mL of BHI and grown shaking at 37°C. Samples were taken at 24 and 48 hour timepoints. Samples were track diluted onto BHI and BHI supplemented with 10 µg/mL of chloramphenicol to calculate colony forming units per mL (CFU/mL). The abundance of the parental strain was calculated by subtracting the CFU/mL on the BHI plate by the CFU/mL on the chloramphenicol plate. Parent measurements which fell below the limit of detection, 1000 CFU/mL, were not included in competitive index calculations. Competitive index was calculated as below and results were summarized using the geometric mean and a 95% confidence interval for each timepoint and ratio^61^.

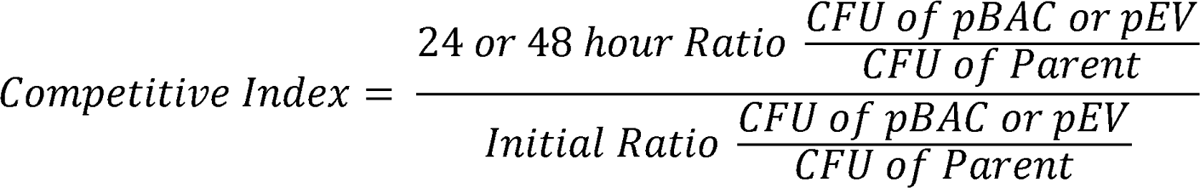

### Mouse Experiments

Animal experiments were approved by the Animal Care and Use Committees of the Children’s Hospital of Philadelphia (IAC 18–001316). Five-week-old C57BL/six male mice were purchased from Jackson Laboratories and given one week to equilibrate their microbiota prior to experimentation. All experimental procedures were performed in a biosafety level two laminar flow hood. Mice were given vancomycin (1mg/mL) in drinking water *ad libitum* for 5 days followed by a 2-day recovery period^62,63^. Mice were then infected with 5×10^8^ *Enterococcus faecium* cells by oral gavage twice a day for two days. Strains were prepared by growing to stationary phase and washing with cold PBS immediately prior to infection. Stool samples were collected daily for quantification of bacterial CFUs. Samples were diluted and homogenized in PBS, and serially plated onto either Bile Esculin Azide (BEA) agar to quantify the total enterococcal population or BEA agar with chloramphenicol (10ug/mL) to quantify the pBAC and pEV strains.

### Global Isolate Collection

All *Enterococcus faecium* genomes deposited in NCBI were downloaded on May 23^rd^, 2024. *E. faecium* genomes with collection dates between 2002-2022 and for which the “host” in the BioSample metadata was listed as “Homo sapiens”, “Homo sapiens sapiens”, “hospitalized patient”, and “Human being” were included. Genomes encoding the *vanA* or *vanB* operon, as identified using AMRFinderPlus, were retained for analysis.

### Statistical Analyses

A single proportion hypothesis test was performed to assess enrichment of cluster isolates within ST groups, setting the proportion of cluster isolates in the total collection as the null value. The association of bacteriocin T8 presence with growth inhibition, liquid competitive advantage of pBAC vs pEV relative to the parent strain, and murine GI tract colonization differences between pBAC and pEV, were assessed using a two-tailed Mann-Whitney test. Bacteriocin T8 enrichment in STs was assessed with a single proportion hypothesis test, using the overall proportion of bacteriocin T8-positive isolates as the null value. Differences in the number of antimicrobial resistance genes, plasmid replicons, and virulence genes between lineages were assessed using one-sided t-tests. Statistical significance was determined with an α = 0.05 and a Bonferroni correction for multiple comparisons was applied when appropriate.

## Code Availability

Not applicable.

## Data Availability

Genomic sequences for all 710 VREfm isolates can be found BioProject PRJNA475751 with accession numbers listed in Supplementary Table 1. All other study data are included in the manuscript and supplementary information.

## Acknowledgements

We gratefully acknowledge Yanhong Li, Kady Waggle, and Hunter Coyle for their contributions to this study, and Dr. Vaughn Cooper and Alecia Rokes for assistance performing whole genome sequencing of EDS-HAT isolates.

## Author Contributions

E.G.M., L.H.H., J.P.Z., and D.V.T. designed the study. E.G.M., A.B.S., M.P.G., K.H., L.P., and A.J.S. performed the experiments and collected data. E.G.M., J.P.Z., and D.V.T. analyzed and interpreted the data. E.G.M. and D.V.T. drafted the manuscript and figures. All authors reviewed and edited the manuscript.

## Ethics Declarations

The authors declare no relevant competing interests.

---

**Figure S1:**
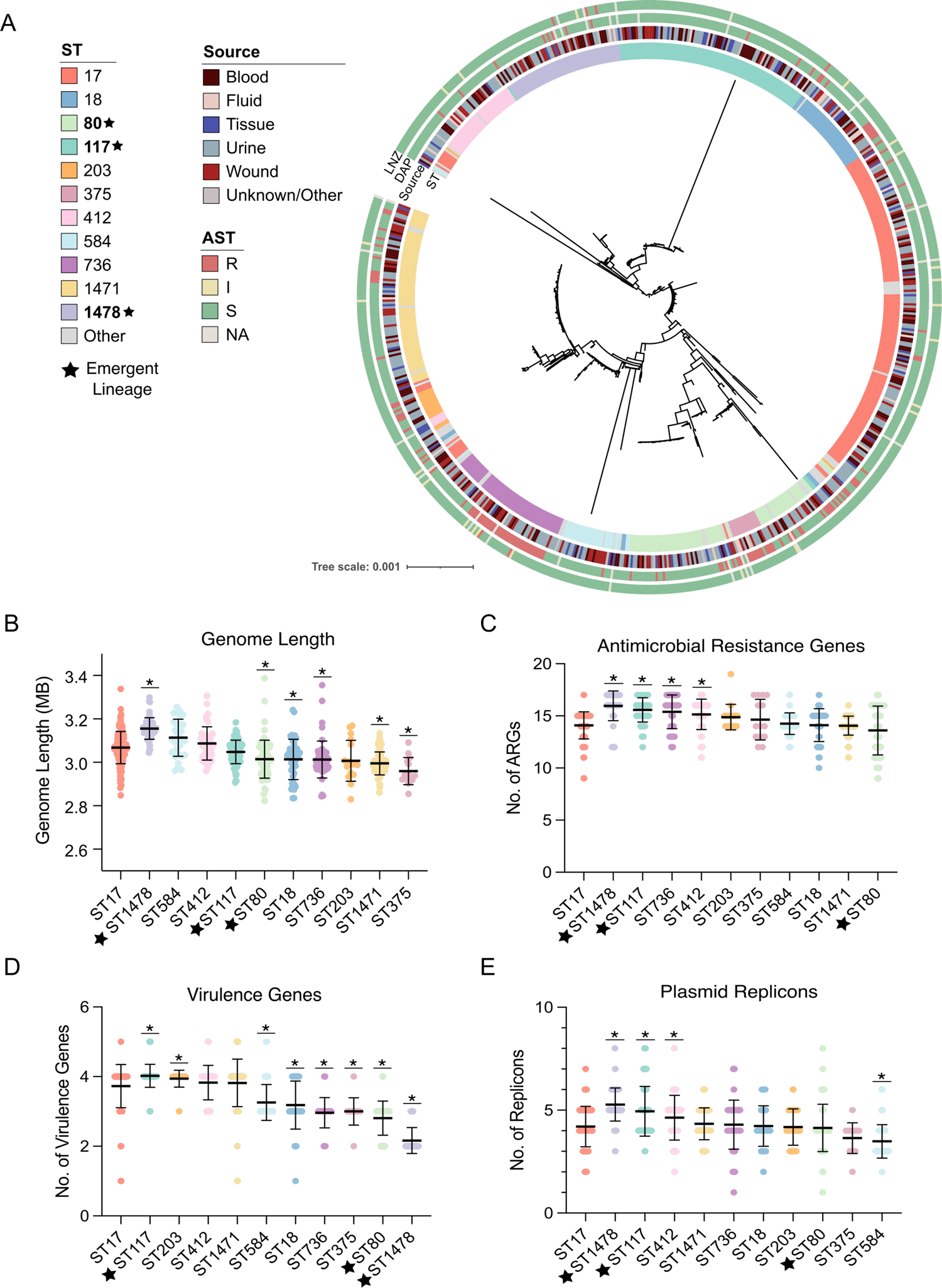
Genetic relatedness and genomic features of 710 VREfm isolates from UPMC. (A) The midpoint-rooted phylogenetic tree was constructed from a core genome alignment. Sequence types (ST) and isolation source are colored as indicated. Emergent lineages are noted by a black star. Antimicrobial susceptibility testing (AST) results for daptomycin (DAP) and linezolid (LNZ) were interpreted as resistant (R), intermediate (I), and susceptible (S). (B) Genome length, (C) presence of antimicrobial resistance genes, (D) presence of virulence genes, and (E) presence of plasmid replicons by main VREfm lineages. Averages of genomic features within each ST were compared to the average seen in the previously dominant ST17 lineage using a one-sided t-test. Asterisks indicate p-values <0.0045. Horizontal lines represent the average and error bars show the standard deviation for each group.

**Figure S2:**
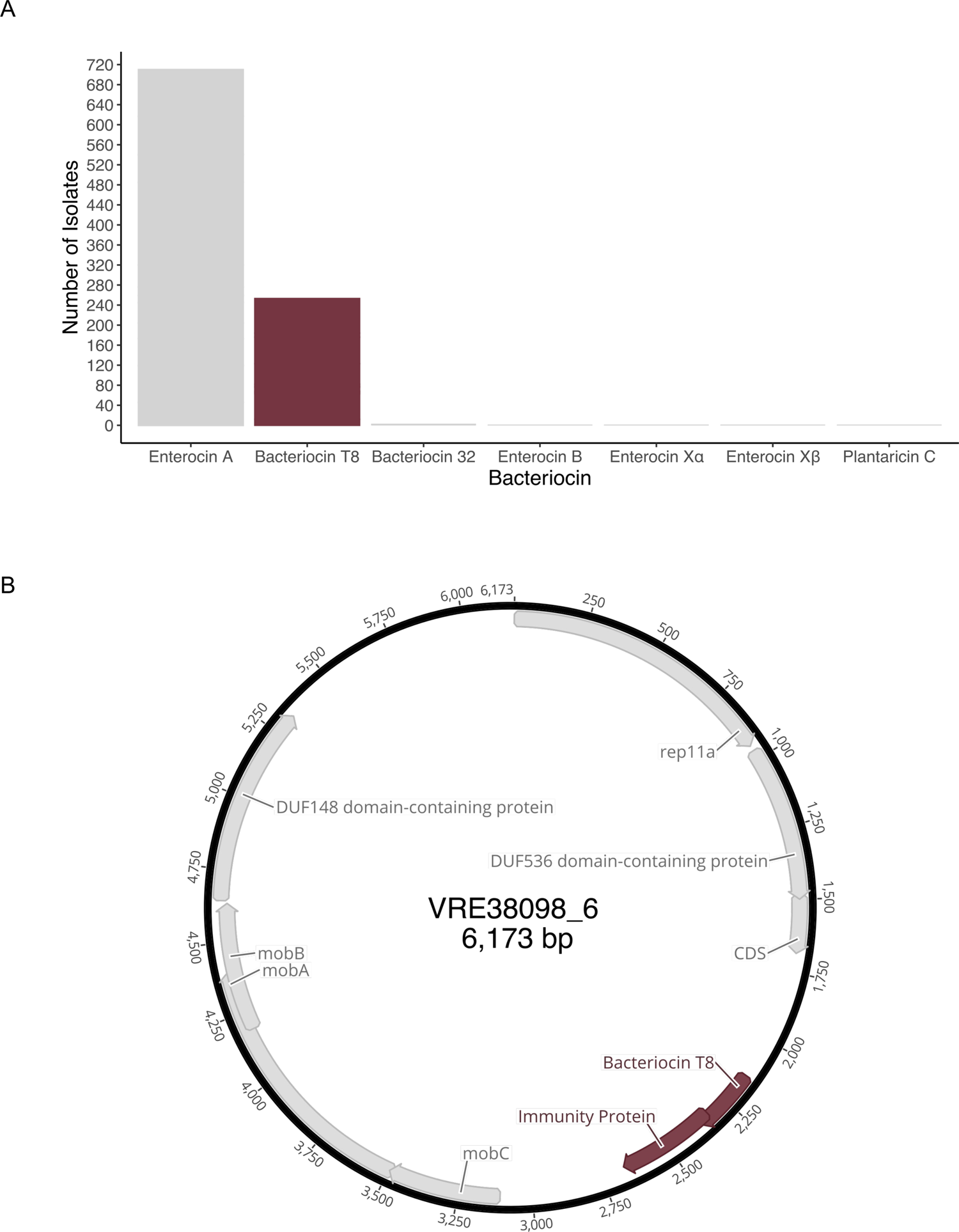
Bacteriocin prevalence and genomic context of bacteriocin T8. (A) Distribution of bacteriocins within 710 VREfm isolates from UPMC. Bacteriocins were identified using BAGEL4 with sequence identity and coverage thresholds of ≥ 95%. (B) Bacteriocin T8-encoding rep11a plasmid from the ST117 isolate VRE38098. Bacteriocin T8 and immunity factor are highlighted in burgundy.

**Figure S3.**
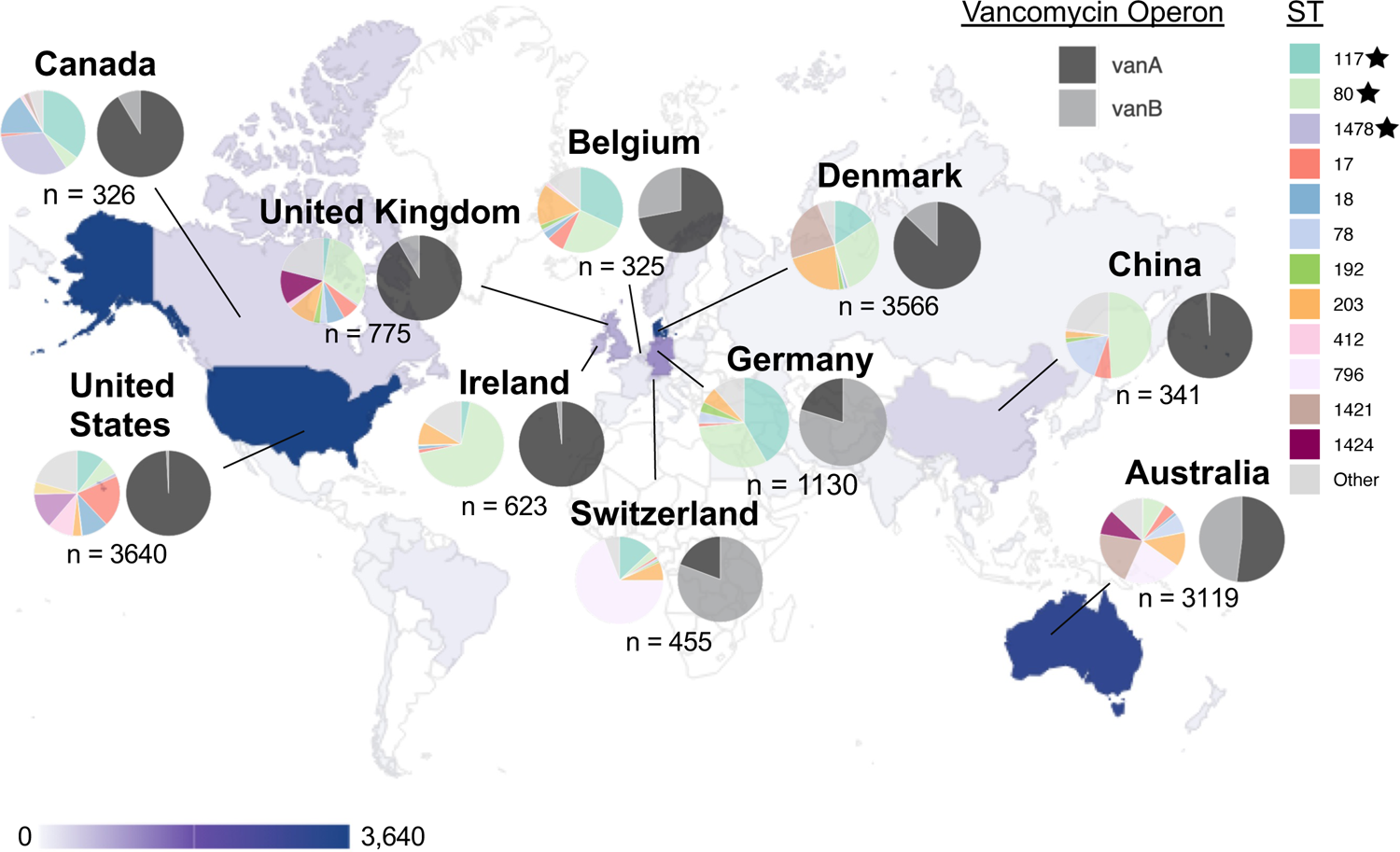
Global population structure of 15,631 VREfm genomes from human sources. Geographical distribution of VREfm genomes pulled from NCBI. The number of genomes from each country is shown from lowest (light grey) to highest (purple). Countries with >300 genomes are highlighted with the distribution of STs and vancomycin resistance operons.

